# Photo-Integrated Conversation Moderated by Robots for Cognitive Health in Older Adults: A Randomized Controlled Trial

**DOI:** 10.1101/19004796

**Authors:** Mihoko Otake-Matsuura, Seiki Tokunaga, Kumi Watanabe, Masato S. Abe, Takuya Sekiguchi, Hikaru Sugimoto, Taishiro Kishimoto, Takashi Kudo

**Author notes:** Corresponding author Mihoko Otake-Matsuura, Ph.D., Team Leader, Center for Advanced Intelligence Project, RIKEN, Nihonbashi 1-chome Mitsui Building, 15th floor, 1-4-1 Nihonbashi, Chuo-ku, Tokyo 103-0027, Japan.

## Abstract

**Background and Objectives:** Social interaction might prevent or delay dementia, but little is known about the specific effects of various social activity interventions on cognition. This study conducted a single-site randomized controlled trial (RCT) of Photo-Integrated Conversation moderated by a Robot (PICMOR), a group conversation intervention program for resilience against cognitive decline and dementia.

**Research Design and Methods:** In the RCT, PICMOR was compared to an unstructured group conversation condition. Sixty-five community-living older adults participated in this study. The intervention was provided once a week for 12 weeks. Primary outcome measures were the cognitive functions; process outcome measures included the linguistic characteristics of speech to estimate interaction quality. Baseline and post-intervention data were collected. PICMOR contains two key features: (i) photos taken by the participants are displayed and discussed sequentially; and (ii) a robotic moderator manages turn-taking to make sure that participants are allocated the same amount of time.

**Results:** Among the primary outcome measures (i.e., cognitive functions), verbal fluency significantly improved in the intervention group. Among the process outcome measures (i.e., linguistic characteristics of speech), the amount of speech and richness of words were larger for the intervention group.

**Discussion and Implications:** This study demonstrated for the first time the positive effects of a robotic social activity intervention on cognitive function in healthy older adults via RCT. The group conversation generated by PICMOR may improve participants’ cognitive function controlling the amount of speech produced to make it equal. PICMOR is available and accessible to community-living older adults.

---

Cognitive health is a key component of healthy aging. Interventions for risk factors may delay or prevent a third of dementia cases (Livingston et al., 2017). While a systematic review found that social activity intervention may help maintain cognitive function among healthy older adults (Kelly, Duff, & Loughrey, 2017), there are no global recommendations for social activity interventions related to cognitive health because evidence of social activity intervention’s impact is limited (World Health Organization [WHO], 2019). Thus, determining the effectiveness of social activity intervention on cognitive health is necessary.

Among the social activity interventions that exist, group-based conversation is the type that is expected to affect cognitive function in particular. Group conversation is a fundamental part of social interaction. Cohort analysis suggests that weekly verbal interactions are associated with verbal learning and memory (Zuelsdorff et al., 2019). Group sessions of cognitive stimulation therapy have also been shown to improve cognition in patients with mild-to-moderate dementia (Woods, Aguirre, Spector, & Orrell, 2012). However, group conversation interventions’ effects on the cognitive functions of healthy older adults is unclear. This is in part because of the difficulty in validating the effects of group conversation, a difficulty resulting from the fact that participation in a group conversation is not easy for older adults who have sensory deficits and/or decrements in language comprehension and production (Gerontological Society of America, 2012). Cognitive changes in older adults are highly variable from person to person, which may also lead to diversity in the level and manner of participation in a group conversation and of outcomes. If there is enough of an imbalance in the amount of speech for the participants, the participants may end up participating in functionally different cognitive tasks. However, no study so far has measured the manner of participation in a group conversation, what conditions regulate it, or its differential effects on cognition in healthy older adults.

Research on technology and aging is growing (Pruchno, 2019), with some studies considering tools and interventions for social connectivity and reduced loneliness (Czaja, Boot, Rogers, Charniss, & Sharit, 2018), well-being (Pu, Moyle, Jones, & Todorovic, 2019), and physical, social, and cognitive activity (Croff, 2019). Inspired by these initiatives, we propose Photo-Integrated Conversation moderated by a Robot (PICMOR), which contains two key technologies: (i) a group conversation support method called Coimagination (Otake, Kato, Takagi, & Asama, 2011; Otake-Matsuura, 2018), in which each participant is allocated an equal amount of time for talking, listening, and question & answer time, and prepares topics and takes photos beforehand according to sessional themes; and (ii) a robot that measures each participant’s speech and supports turn-taking on that basis during the discussion phase of the intervention (Yamaguchi, Ota, & Otake, 2012). The Coimagination method follows the recommendations of the Gerontological Society of America (2012), which are oriented to eliminate older adults’ communication difficulties with healthcare professionals, but they are applicable to the communication among older adults as well. Specific recommendations are the use of photos as supports, giving each participant equal opportunity to talk, and verifying comprehension through question and answer sessions.

This study’s purpose was to gather evidence of the effects of PICMOR on cognition in healthy older adults, and to validate PICMOR using a randomized controlled trial (RCT). We will discuss the effects and their possible sources. Group conversation without guidance or feedback was used in the control group; conversation was encouraged in both groups (instead of using a control group with less conversation) to allow variation in speech amounts among participants to emerge and examine the possibility that balanced speech may have positive effects on the cognition of older adults, in particular, verbal production and comprehension.

This paper first focuses on the primary and secondary outcome measures of the trial, that is, cognitive functions and quality of life. Then, we explore process outcome measures: linguistic characteristics of speeches in both groups to compare interaction quality, and the number of photos taken and memory recall scores of the intervention to estimate engagement. Our hypothesis is that participants who complete the PICMOR intervention will show subsequent improvement on certain subcategories of cognitive functions and quality of life from baseline to post-treatment compared to participants in the active control program.

## Method

### Study Design and Procedures

The 12-week RCT took place from June to September 2018 (UMIN Clinical Trials Registry number: UMIN000036667). Figure 1 presents the CONSORT flowchart of this study. First, screening and baseline assessment (medical interviews, neuropsychological tests, and self-reported questionnaires) were conducted to determine participant eligibility. After baseline assessment, participants were randomly assigned to the intervention or control group according to the Japanese version of the Mini-Mental State Examination (MMSE-J) scores (Sugishita et al., 2018) and Japanese version of Montreal Cognitive Assessment (MoCA-J) scores (Fujiwara et al., 2010). After 12 weeks of intervention, a post-assessment was conducted. There were no drop-outs to follow-up during the intervention. The assessors were not involved in the intervention delivery.

**Figure 1.**
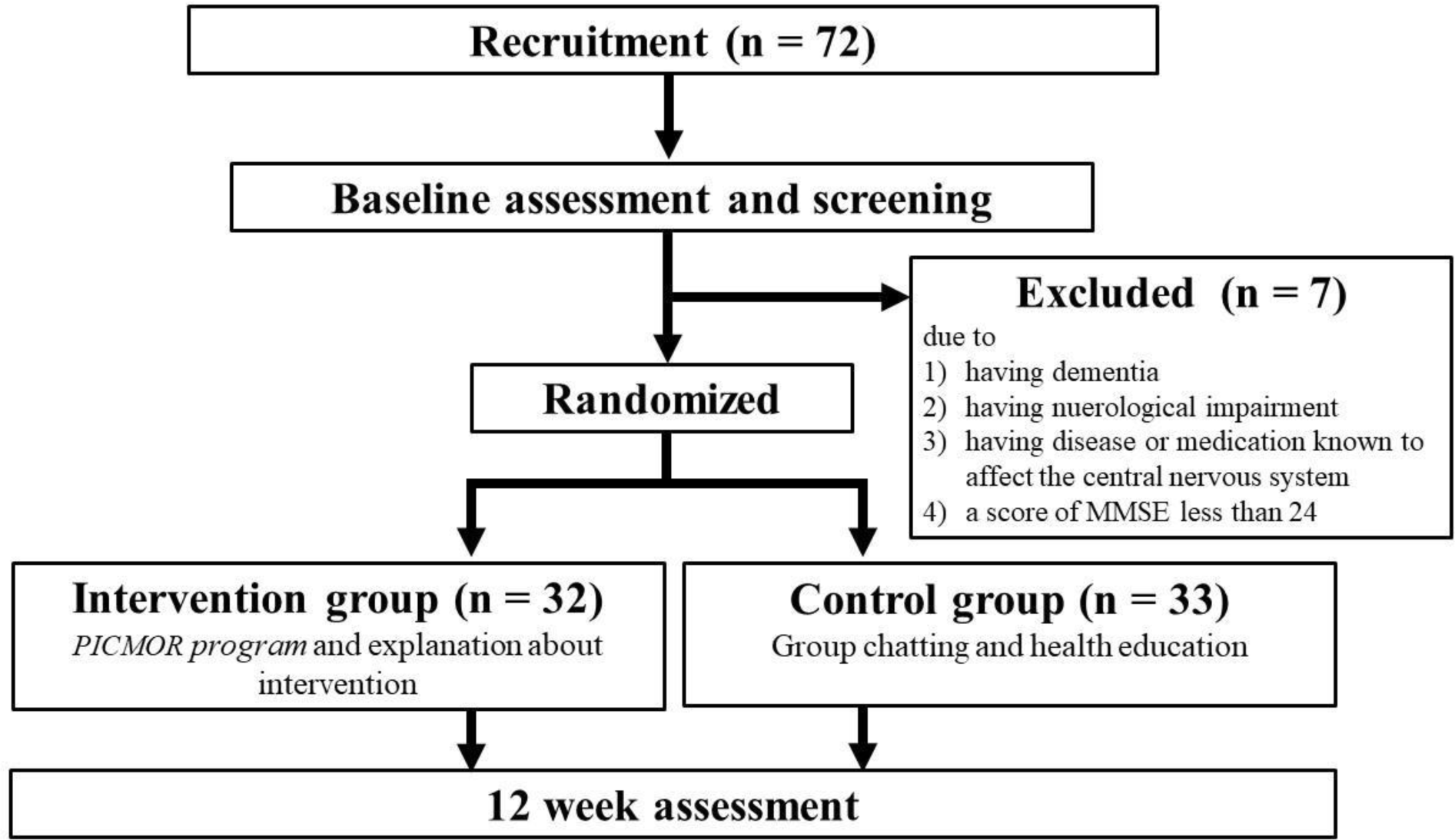
CONSORT diagram flowchart.

Intervention group participants received weekly 30-minute intervention sessions, each followed by 30 minutes of explanation about the intervention. The active control sessions in the control group involved 30 minutes of weekly unstructured conversation among the group and 30 minutes of health education about successful aging. Each group was divided into four-person subgroups with both men and women, formed on the basis of participants’ availability.

The Institutional Review Board approved this study. All participants provided written informed consent.

### Participants

The participants were community-living healthy adults aged over 65 years, recruited from the Silver Human Resources Center. The exclusion criteria were as follows: dementia; neurological impairment; any disease or medication known to affect the central nervous system; MMSE-J score less than 24. Seventy-two people received screening.

### Intervention: PICMOR

PICMOR is an integrative intervention program supporting the preparation of conversation topics, time management, and turn-taking in conversations, and reflection on the topics. The PICMOR program consists of three phases: preparation, conversation, and reflection (see Figure 2.) The main phase is conversation. In order to make participants well-prepared and focused during the conversation, preparation and reflection phases precede and follow the conversation, respectively.

**Figure 2.**
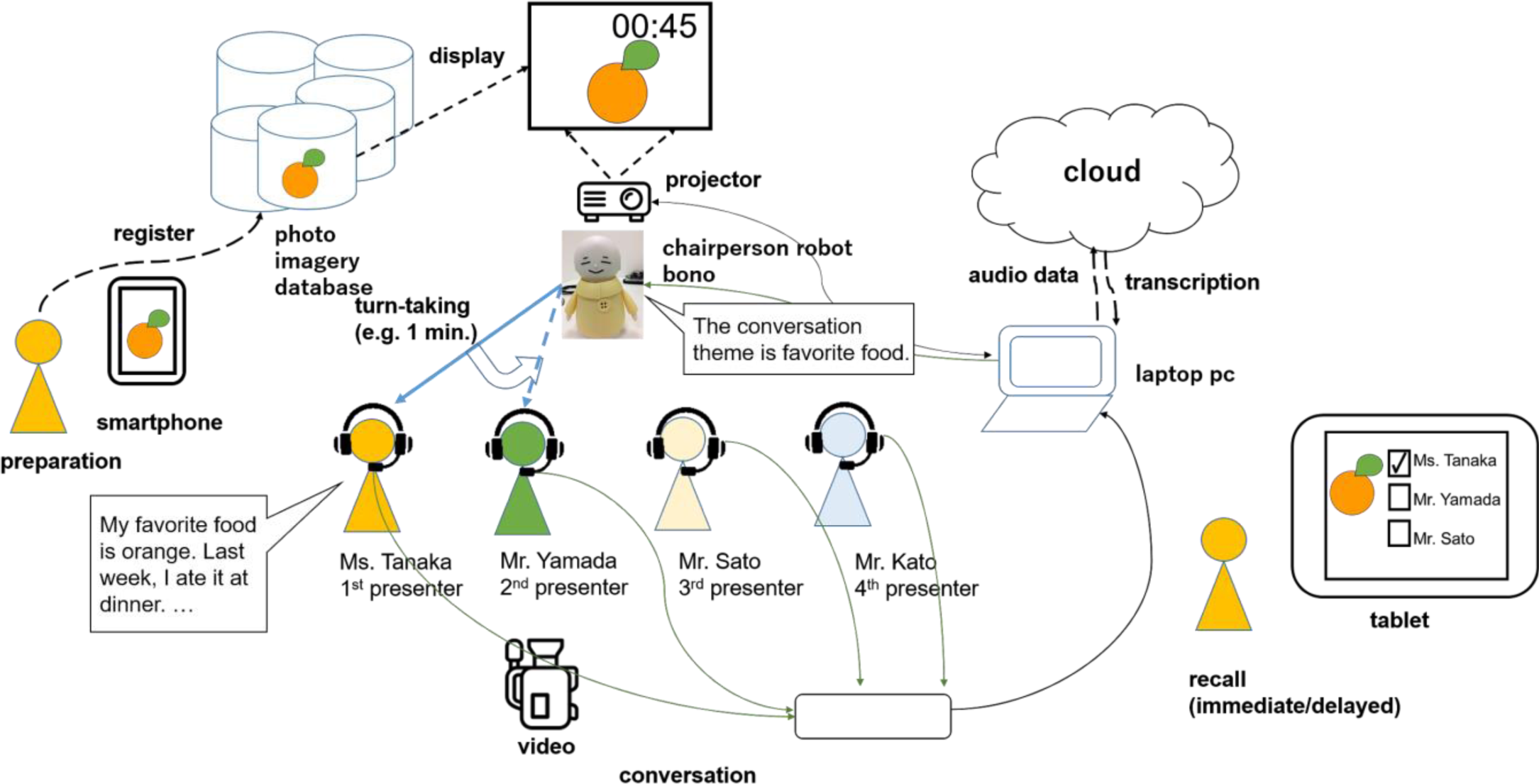
Experimental setup for Photo-Integrated Conversation Moderated by a Robot (PICMOR).

#### Preparation Phase

During this first phase, each participant used a smartphone with a specially developed application to take photos that represent topics related to a theme. We tested the application beforehand to ensure that older adults using smartphones for the first time would be comfortable. The initial screen of the application comprised only two buttons: one to take a photo, and another to select photos for conversation. Photos were smoothly uploaded to the online PICMOR database. The participants’ assignment was to take photos and find topics based on a set theme of the week for participants to talk about. When the theme of the session was “Favorite Food,” for example, the topic of the first presenter, Ms. Tanaka, was “Orange.” The theme is announced before a given week’s session (see the left of Figure 2). Since the program was provided weekly, the theme for the following week’s session was announced to participants at the end of each session. In this study, each participant engaged with 12 themes because he/she has participated for 12 weeks in the experiment. The 12 themes included favorite places in the neighborhood, tips for health, and comparison of before and after cleaning. The themes were designed to trigger activities that produce new episodic memories. The location, timing, and frequency of taking photos depended on the participant. They were asked to take as many photos as possible and select the best two. In total, 24 photos were used for each participant during the 12 weeks of intervention.

#### Moderated Conversation Phase

In the second phase, participants are cued to talk when their photos are displayed on the screen. Each photo is displayed one after another as a set, and each set is presented twice. In the first round, all participants describe their own photos. In the second round, all participants discuss each other’s photos (see the center of Figure 2, where participants are in a conversation setting using the PICMOR method, looking at their photos). Participants were divided into groups of four people. The conversation session consisted of two stages. First, each participant was assigned two minutes to present two photos related to the topic, each of which was displayed for one minute. Second, during the discussion, the other participants asked questions and gave comments to the presenter. Each participant was assigned four minutes for discussion, during which each photo was displayed for two minutes. This process was repeated for each participant. Each session lasted about 30 minutes, including the instructions given by a robot and the interval between each presentation and discussion.

Conversations were moderated by a “chair-robot” developed by us. Named Bono, this robot is proficient at time management. A robot was chosen instead of a human because a robot has skills such as turn-taking moderation based on analysis of speech patterns that a human does not. Bono has four degrees-of-freedom — head pitching, body rotation, and left and right arm elevations — which are sufficient for its moderator role. For instance, when a speaker finishes talking, the robot is able to select the next speaker by rotating its body at a given angle and lifting its right arm horizontally to address that person directly. With a loud speaker installed inside of the main body of the robot, Bono can say to a participant, “Please talk about your photo.” In this study, Bono automatically and strictly moderated the conversation and conducted turn-taking based on the time slot duration previously determined by the researchers. Followings are robot’s skills a human moderator does not possess: During the discussion, the robot would prompt and stop participants’ utterances automatically based on total speech time, silent time, and utterance length so as to balance the production of speech for each participant (Yamaguchi et al., 2012); When the robot detected that a participant had spent less time than the others on conversation, it would directly encourage the participant to provide questions or comments. Each participant wore a headset-microphone that recorded his/her voice to measure each participant’s speech precisely. This audio data was transmitted in real-time from the microphones to a computer via cables to precisely measure and balance the amounts of speech and to transcribe the speech for linguistic analysis. In this study, we also recorded videos to capture the details of each conversation. Participants were filmed from behind to protect their privacy.

#### Recall Phase

In the last phase, the participants completed memory tasks using a specially developed tablet application. The photos previously displayed during the conversation were randomly shown, and the participants were asked to indicate the presenter who took the photo by touching the name on the touch panel. These tasks were conducted at two points in time: soon after the end of the conversation in order to measure immediate recall and one week later in order to measure delayed recall. The immediate recall task was meant to check if the participants could focus on listening and understanding, resulting in remembering the presenter’s name of each photo. The delayed recall task was conducted just before the next conversation of the week to check whether the participant’s memories were preserved.

### Outcome Measures

The primary outcome measures in the present study were cognitive performance measures evaluated by standardized neuropsychological tests conducted by well-trained examiners.

The MMSE-J (Sugishita et al., 2018.) and MoCA-J (Fujiwara et al., 2010) were administered to evaluate global cognitive function. The logical memory subtests Logical Memory I and II from the Wechsler Memory Scale-Revised (WMS-R) (Wechsler, 1987) were introduced to evaluate memory function. Logical Memory I assesses immediate recall of the content of a story immediately after the examiner reads it, while Logical memory II assesses delayed recall 30 minutes later. Two kinds of stories were used, one chosen randomly at base assessment and another used at post-assessment. The Advanced Trail Making Test (ATMT) (Mizuno & Watanabe, 2008) assesses attention and executive functions using a computer. The Wechsler Adult Intelligence Scale — Third Edition (WAIS-iii) (Wechsler, 1997), Digit Span Forward and Backward, and Digit Symbol Coding tests were also used. Digit Span Forward assesses simple memory span, and Digit Span Backward assesses working memory capacity. The Digit Symbol Coding test assesses the process speed and memory in digit symbol coding performance, which requires the subject to write down each corresponding symbol as fast as possible. In the verbal fluency tests, letter fluency was evaluated to measure verbal function; specifically, participants were asked to pronounce as many words as possible starting with the Japanese character “ka” in one minute, and then the total number of words was counted.

The secondary outcome measures covered subjective physical and mental status and quality of life. The Tokyo Metropolitan Institute of Gerontology-Index of Competence (TMIG-IC) (Koyano, Shibata, Nakazato, Haga, & Suyama, 1991), the Japanese version of the Geriatric Depression Scale short form (GDS-15-J) (Sugishita, Sugishita, Hemmi, Asada, & Tanigawa, 2017), and the WHO Quality of Life questionnaire 26 (WHO QOL 26) (Tazaki & Nakane, 1997) were used.

The process outcome measures were the linguistic characteristics of speeches in both groups to compare the quality of interactions, and the number of photos taken and memory recall scores to estimate engagement in the intervention group.

### Analysis

For basic characteristics at baseline, Welch’s t-test was used to compare the means of continuous variables (Age, MMSE-J, MoCA-J, GDS-15, and TMIC-IC) and Fisher’s exact test were used to compare frequency distributions of categorical variables (Gender and Education) between groups.

To estimate the intervention effects on the aforementioned outcome measures, linear mixed models with random-effect intercepts for participants were performed for all outcome measures using the “lmer” function in the R package, “lme4” (Bates, Mächler, Bolker, & Walker, 2015). The models have the following independent variables: *time* (1 = post-experiment; 0 = pre-experiment), *group* (1 = intervention group; 0 = control group), and their interaction term *time*× *group*, which is interpreted as the intervention effect. To obtain p-values associated with the linear mixed analyses, we used the R package “lmerTest” (Kuznetsova, Brockhoff, & Christensen, 2017). It was confirmed visually that there were no severe deviations from straight lines in normal quantile–quantile plots. Taking the relatively small sample size into account, we judged that the assumption of normality was reasonably valid. However, for TMIG-IC, TMTs-A and −B, large deviations from straight lines were observed. Therefore, for TMTs-A and −B, the mixed linear models were performed for logarithmic transformed outcomes. For TMIG-IC, instead of using the mixed linear model, we performed the mixed Poisson regression analysis for the number of items indicating lower functional capacity, by using the “glmer” function in the R package, “lme4” (Bates et al., 2015).

### Analysis of conversation

To investigate differences in conversation patterns between the control and intervention groups, we quantitatively analyzed conversation transcriptions derived from the audio data. We focused on the number of words and lexical richness (i.e., type–token ratio) after decomposing the sentences into words, rather than the meanings of conversations or words. The reason is that when we consider the cognitive function in conversation, how much information participants can express through conversation is considered important (Snowdon et al., 1995; Kemper, Thompson, & Marquis, 2001). Therefore, we quantified the conversational characteristics for each participant as a simple index based on the number of unique words.

For the analysis, first, we used Google Cloud Speech-to-Text (Google, Mountain View, CA, 2018) to automatically transcribe audio to text data, and then manually checked the entire text by comparing it to the audio data and fixing any mistakes. Second, we conducted morphological analysis using “MeCab” (ver. 0.996), a useful tool for Japanese morphological analysis based on conditional random fields (Kudo et al. 2004). Finally, we calculated the number of spoken words per time unit (per minute), the standard deviation of the number of spoken words in each session, and lexical richness for each participant. We used bilogarithmic type-token ratio (logTTR), defined as log(number of types)/log(number of tokens), to quantify lexical richness (Wachal & Spreen, 1973). If this value is high, the speech contains much more information in terms of the number of vocabulary items. We conducted these analyses with R Ver. 3.4.3 (R-Core Team, Vienna, Austria, 2017).

#### Analysis of photos

We evaluated participants’ engagement in preparing topics for future conversations based on the number of photos taken per session per participant, under the assumption that more photos indicated more effort to find topics for the next conversation session. We also referred to the participants’ comments on why they took a large or small number of photos.

We evaluated the accuracy rates of the photo recall tasks. If the accuracy rate of the immediate recall task was high, we assumed that attention and short-term memory were functioning well. Similarly, if the accuracy rate of the delayed recall task was high, recent memory was estimated to be well-functioning.

## Results

Seven participants were excluded at screening because they meet our exclusion criteria. Therefore, a sample of 65 people was divided into intervention and control groups (intervention: *n*=32, control: *n*=33). Participants in each group were divided into 8 subgroups of 4–5 participants. Basic characteristics of study participants at baseline are presented in Table 1. The only significant difference found between groups was a significantly higher GDS-15 score in the control group. All participants completed the program and post-measurement.

**Table 1.** Baseline Group.

|  | Intervention<br>(*N*=32) | Control<br>(*N*=33) | *p* |
| --- | --- | --- | --- |
| Age (Mean ± SD) | 72.97±3.47 | 72.33±2.90 | 0.427 |
| Gender (Female; *n*, %) | 16 (50.0%) | 19 (57.6%) | 0.622 |
| Education (≥13 years; *n*, %) | 20 (62.5%) | 17 (51.5%) | 0.455 |
| MMSE-J (Mean ± SD) | 27.97±1.43 | 28.12±1.52 | 0.677 |
| MOCA-J (Mean ± SD) | 25.97±2.47 | 25.42±2.66 | 0.395 |
| GDS-15 (Mean ± SD) | 2.09±1.73 | 3.36±2.18 | 0.011* |
| TMIG-IC |  |  |  |
| Total score (Mean ± SD) | 12.31±1.03 | 11.88±1.32 | 0.144 |
| The instrumental activity of daily living (Mean ± SD) | 5.00±0.00 | 4.97±0.17 | 0.325 |
| Intellectual activity (Mean ± SD) | 3.84±0.51 | 3.67±0.74 | 0.265 |
| Social role (Mean ± SD) | 3.47±0.67 | 3.24±0.83 | 0.231 |

*Note.* Welch's t-tests for Age, MMSE-J, MoCA-J, GDS-15, and TMIG-IC Fisher's exact test for Gender and Education. Abbreviations. MMSE-J: Japanese version of the Mini-Mental State Examination, Japanese version; MoCA-J: Japanese version of the Montreal Cognitive Assessment; GDS-15: 15 - item Geriatric Depression Scale; TMIG-IC: Tokyo Metropolitan Institute of Gerontology Index of Competence; SD: standard deviation.
\**p*<0.05

### Primary and Secondary Outcomes

Table 2 summarizes cognitive test scores within participants (pre- and post-experiment) and between participants (intervention and control groups). In Logical Memory I and II and Digit Symbol tests, overall scores significantly improved after the intervention. However, there was no significant *time* × *group* interaction on the scores. Regarding MMSE-J, MoCA-J, forward and backward Digit Spans, and TMTs-A and −B, neither main effects nor interaction effects were found. In the verbal fluency test, a significant *time* × *group* interaction was obtained. The regression coefficient of *time* × *group* associated with verbal fluency was 2.024, meaning the number of generated words between pre- and post-experiment was approximately two words more than in the control group—from 11.8 at pre-experiment to 13.6 at post-experiment—while there was little change in the control group from pre-experiment (11.4) to post-experiment (11.2). In all secondary outcomes—TMIG-IC, GDS-15, and WHO QOL26—no intervention effects were found.

**Table 2.** Comparison of Pre/Post Cognitive Test Scores in Intervention and Control Groups.

|  | Intervention (*n*=32) |  | Control (*n*=33) |  | Estimates |  |  |
| --- | --- | --- | --- | --- | --- | --- | --- |
|  | Pre Mean<br>(*SD*) | Post Mean<br>(*SD*) | Pre Mean<br>(*SD*) | Post Mean<br>(*SD*) | Time<br>(SE, *p*) | Group<br>(SE, *p*) | Intervention<br>(SE, *p*) |
| MMSE-J | 28.0<br>(1.4) | 28.6<br>(1.8) | 28.1<br>(1.5) | 28.7<br>(1.2) | 0.545<br>(0.30, 0.074) | -0.152<br>(0.37, 0.683) | 0.111<br>(0.43, 0.797) |
| MoCA-J | 26.0<br>(2.5) | 26.3<br>(2.7) | 25.4<br>(2.7) | 25.2<br>(2.9) | -0.212<br>(0.44, 0.631) | 0.545<br>(0.66, 0.413) | 0.556<br>(0.63, 0.378) |
| Logical memory I<br>(immediate) | 10.2<br>(3.2) | 11.3<br>(3.5) | 8.3<br>(4.0) | 10.8<br>(4.2) | 2.515***<br>(0.72, 0.001) | 1.915*<br>(0.93, 0.042) | -1.39<br>(1.02, 0.178) |
| Logical memory II<br>(delayed) | 8.5<br>(3.0) | 9.5<br>(3.5) | 6.5<br>(3.6) | 9.1<br>(4.3) | 2.606***<br>(0.66, 0.000) | 2.016*<br>(0.90, 0.028) | -1.606<br>(0.93, 0.091) |
| Verbal fluency | 11.8<br>(3.7) | 13.6<br>(3.5) | 11.4<br>(4.3) | 11.2<br>(3.7) | -0.242<br>(0.60, 0.686) | 0.450<br>(0.95, 0.637) | 2.024*<br>(0.85, 0.020) |
| Symbol<sup>1)</sup> | 55.2<br>(13.1) | 66.5<br>(11.9) | 52.0<br>(13.7) | 61.7<br>(11.4) | 9.667***<br>(2.40, 0.000) | 3.194<br>(3.14, 0.312) | 1.624<br>(3.45, 0.640) |
| Digit Span Forward | 9.8<br>(2.0) | 9.7<br>(1.6) | 9.6<br>(1.7) | 10.1<br>(1.6) | 0.455<br>(0.26, 0.085) | 0.114<br>(0.44, 0.796) | -0.517<br>(0.37, 0.167) |
| Digit Span Backward | 6.4<br>(1.7) | 6.4<br>(1.9) | 6.1<br>(1.5) | 6.5<br>(1.5) | 0.394<br>(0.28, 0.165) | 0.347<br>(0.41, 0.399) | -0.394<br>(0.40, 0.329) |
| TMT-A<sup>2)</sup> | 76.1<br>(36.2) | 65.5<br>(21.1) | 90.5<br>(58.3) | 87.6<br>(64.5) | -0.031<sup>3)</sup><br>(0.04, 0.448) | -0.105<sup>3)</sup><br>(0.11, 0.346) | -0.077<sup>3)</sup><br>(0.06, 0.184) |
| TMT-B<sup>2)</sup> | 141.5<br>(107.5) | 103.5<br>(42.3) | 140.9<br>(88.5) | 121.3<br>(61.9) | -0.099<sup>3)</sup><br>(0.07, 0.133) | -0.012<sup>3)</sup><br>(0.12, 0.918) | -0.113<sup>3)</sup><br>(0.09, 0.232) |

<sup>1)</sup> One participant with a missing value in the intervention group was eliminated.
<sup>2)</sup> One participant with a missing value in the intervention group was eliminated.
<sup>3)</sup> Estimations for logarithmic transformed response variables.
Abbreviations. MMSE-J: Japanese version of the Mini-Mental State Examination, Japanese version; MoCA-J: Japanese version of the Montreal Cognitive Assessment; TMT: Trail Making Test.
\**p*<0.05; \*\*\**p*<0.001.

### Process Outcomes

We quantified conversations based on the number of words and lexical richness of the speech and thereby revealed the difference between control and intervention groups. Figure 3(A) shows the number of words per minute. GLMM (log link function, offset as time duration, with random effects of id and group) analysis reveals no significant difference between control and intervention in the number of words (*p* = 0.51). To investigate evenness in the number of words within groups in each session, we calculated the standard deviation of the number of words in each session (Figure 3 (B)). The GLMM (gamma link function, random effect of group) reveals that the standard deviation in the intervention group was smaller than in the control group (*p* = 2.27 ×10^−8^), indicating that participants in the intervention group spoke more evenly than in the control group. Figure 3 (C) shows lexical richness for each participant. The linear mixed model (random effect of group) suggests that logTTR in the intervention group was larger than in the control group (*p* = 0.029). The results demonstrate that the program intervention led the speech to contain more diverse information. Moreover, robot Bono can promote and suppress speech during the session. The average number of instances of promotion and suppression of speech by the robot during each session per participant was 1.20 (*SD* = 1.81) and 0.95 (*SD =* 1.67), respectively.

**Figure 3.**
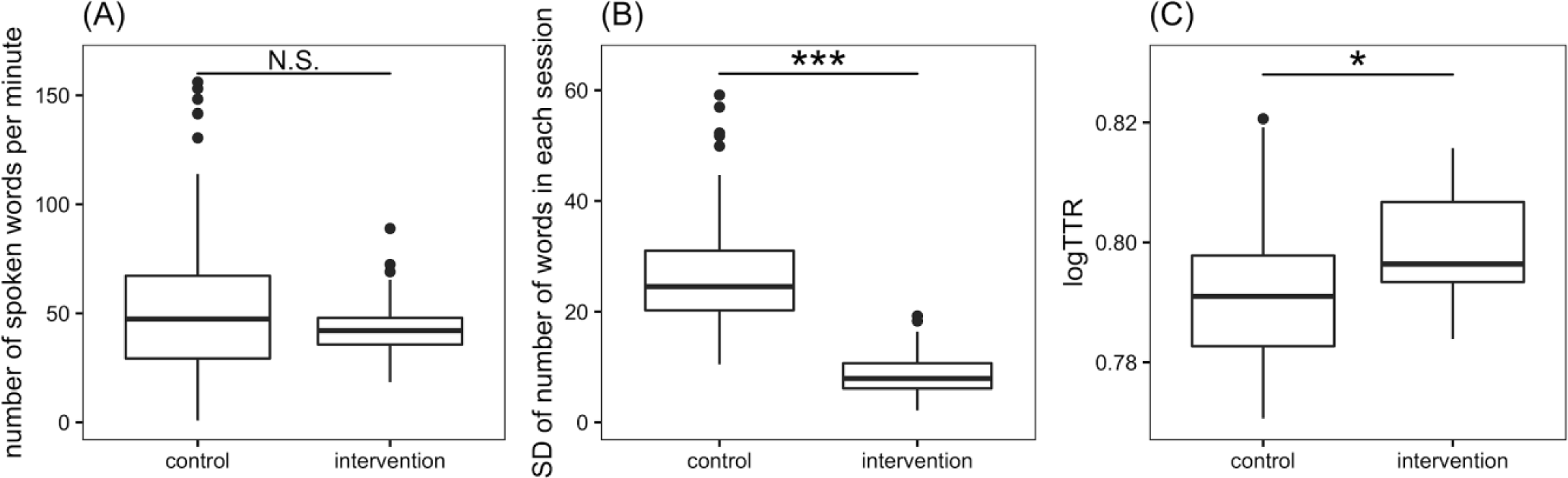
Boxplots for results of conversational analysis. (A) The number of spoken words per minute; (B) standard deviation (*SD*) of number of words; and (C) logTTR: bilogarithmic type-token ratio. \**p*<0.05, \*\*\**p*<0.001.

All participants in the intervention group successfully used smartphones to take photos, although for 62.5% of them, it was their first time using a smartphone. The number of photos per session per participant ranged from 2 to 116 (*M* = 15.43, *SD =* 14.74). Participants who took many photos mentioned that they were interested in or excited about taking photos, while those who took few photos said that they were busy. The total number of photos taken was 5,924. The average scores on immediate and delayed recall tasks were 98.88% and 97.60%, respectively.

## Discussion

While effective interventions that improve cognitive health in aging populations are increasingly necessary (WHO, 2019), evidence regarding the impact of social activity interventions on cognitive functioning remains limited. This study advances contemporary knowledge regarding the impact of social activity intervention in helping older adults participate in group conversation equally. Through a RCT, this study is one of the first to demonstrate the positive effects of social activity intervention – a group conversation regulated by a robot – on cognitive function in healthy older adults.

This RCT examined cognitive functions as primary outcomes for both the control and experimental groups, linguistic characteristics of speech as process outcomes for both groups, and the numbers of photos and memory task scores as process outcomes for the intervention group. The results showed that verbal fluency improved significantly for the intervention group compared to the control group. Previous studies have reported that conversation-based interventions with trained interviewers impacted positively on verbal fluency performance (Cerino, Hooker, & Dodge, 2019; Dodge, Zhu,& Kaye, 2015). In the current study, both inter- and intra-personal variation in the amount of speech was smaller for the intervention group, and their unique word rate was higher than in the control group. This study adds new evidence that providing group conversation guidance comprised of turn-taking support by a robot, displaying photos, and giving feedback is effective to balanced speech amounts across participants so as to improve verbal fluency performance.

### Verbal Fluency

Verbal fluency—more precisely, letter fluency, or the ability to produce words starting with a certain letter—improved for the intervention group compared to the control group. Tests of verbal fluency discriminate well between people with normal cognitive function and mild Alzheimer’s disease (Henry, Crawford, & Phillips, 2004) and are, therefore, used for preliminary diagnosis of dementia in clinical settings. Verbal fluency draws on both executive functions and language abilities. Although verbal fluency declines with age, the slower processing masks the enhancement of letter fluency during the transition from youth to middle age. In one study, an older group performed better than a younger group on letter fluency after controlling for the decline in processing speed (Elgamal, Roy, & Sharratt, 2011). This implies that verbal fluency is a trainable ability that should improve through the life course, and such improvement was indeed observed through the relatively short-term intervention in this study. The theory of cognitive reserve, where brain reserve is related to either the brain’s anatomical substrate or adaptability of cognition (Stern, 2013; Amieva et al., 2014), suggests that more brain reserve helps people tolerate more neuropathology without cognitive or functional decline, and therefore they develop dementia more slowly than do people without brain reserve (Stern, 2012). Improvement of verbal fluency, which declines significantly at the onset of dementia, should increase cognitive resilience. Further, cognitive resilience in later life is likely enhanced by building brain reserve earlier in life through education and other intellectual stimulation (Borenstein & Mortimer, 2016; Larson, 2012). The results of this study suggest that intensive group conversation may serve as one such intellectual activity stimulating cognitive resilience.

### Linguistic Characteristics of Conversation

Linguistic ability is significantly associated with cognitive functions (Kemper et al., 2001; Snowdon et al., 1995). Our results show significant differences in evenness and the amount of information in conversations between the intervention and control groups even while the number of words spoken did not differ (Figure 3). This indicates that PICMOR can both induce participants to speak more and include much more information in their speech while keeping the amount of speech constant. Evenness in speech-sharing is important for conversation as intervention, as it entails balanced use of verbal comprehension and verbal production. In our study, some participants in free conversation (i.e., the control group) tended to speak much more and others less (Figure 3 (B)), suggesting that temperamentally more talkative participants gain more skills at verbal production than verbal comprehension, and vice versa. In our intervention program, all participants had the opportunity to engage in verbal comprehension (listening) and verbal production (speaking), and the robot prompted participants who had had less speech and suppressed those with more speech, fostering evenness in conversation and possibly the higher verbal fluency scores. Moreover, the type–token ratio in the intervention group was higher than that in the control group. This is because participants in the intervention group took photos and made a presentation about things related to the photos, and then discussed them with each other. Furthermore, these participants would compose sentences effectively to pack more information into their speech. Previous studies used “idea density” as a similar index representing the amount of information in sentences and predicting cognitive function (Snowdon et al., 1995). Likewise, this intervention may increase verbal fluency.

### Comparison of Intervention and Control Groups

The scores of both groups improved significantly after the Logical Memory I and II and Digit Symbol tests. The major reason may be that both intervention and control conditions included active group conversation. Another reason may be that the tests had learning effects. The trial will be conducted with a less active control condition, without group conversation, to clarify the main reason for these improvements.

### Applications

PICMOR is applicable to practice and measurement support using methods with group sessions, such as cognitive stimulation therapy and group reminiscence therapy. Methods with group sessions generally require at least one trained facilitator per group, leading to increased training and hiring costs, that is, a scalability problem. PICMOR may increase scalability by obviating the need for human facilitator per group, at least for healthy older adults. A human instructor can remain to support participants who have special needs. With more robots, several group conversations can be coordinated by one human instructor operating multiple robots. In addition, building robots should also become cheaper at increased scale.

### Limitations and Future Work

This study was of relatively short length and held infrequent (i.e., weekly) sessions. Thorough investigation of the demonstrated effect of PICMOR warrants a longer study with more frequent sessions: for example, two or three times a week for two years, as in FINGER, the multi-modal lifestyle intervention study (Ngandu et al., 2015). This should also increase the visibility of the effects. A follow-up study is planned to investigate whether PICMOR may slow down cognitive decline and delay dementia for years. While the verbal function improvement is certainly an effect of conversation, some of the effects of PICMOR may be caused not by group conversation but by using photos and robots. The purpose of this study is to propose an effective, efficient, and reproducible intervention program in which group conversation characterized by balanced speech production is realized. The effect of each item should be further studied to make clear which aspect of PICMOR is essential for positive effects to occur.

## Data Availability

Joint research agreement is required for data sharing.

## Acknowledgments

We are deeply grateful to Yoshinori Fujiwara (Tokyo Metropolitan Geriatric Hospital and Institute of Gerontology), Hiroyuki Suzuki (Tokyo Metropolitan Geriatric Hospital and Institute of Gerontology), Kei Mizuno (RIKEN), Akihiro Sasaki (RIKEN), and all study participants and staff.

